# Secondhand tobacco exposure assessed using urinary cotinine among 10 years children in Japan: A 11-years repeated cross-sectional study

**DOI:** 10.1101/2024.04.25.24306399

**Authors:** Yudai Tamada, Kenji Takeuchi, Takahiro Tabuchi

## Abstract

**Introduction:** The emergence of heated tobacco products (HTPs) has complicated the issue of secondhand tobacco (SHT) exposure. We investigated the trend in SHT exposure in schoolchildren and assessed whether SHT exposure depends on household tobacco use status.

**Methods:** This repeated cross-sectional study from 2011 to 2021 (15,927 subjects were eligible) was based on data from an annual survey of fourth-grade students (aged 10 years) in Kumagaya City, located in Kanto Region, Japan. Data were collected by questionnaire, and urinary cotinine levels used as an objective indicator of SHT exposure, such as secondhand exposure to combustible cigarette (CC) smoke and HTP-generated aerosols.

**Results:** SHT exposure has decreased by about one-quarter over the 11-year period, from 18.6% in 2011 to 5.3% in 2021. Regarding household tobacco product use, 68.1% did not use tobacco products, 16.5% were CC-only users, 12.3% were HTP-only users, and 3.0% were both CC and HTP users. The prevalence of SHT exposure among participants was significantly higher compared to participants with no tobacco product users in the household, in the following order: both CC and HTP users, only CC users, and only HTP user.

**Conclusions:** While the prevalence of SHT exposure showed a decreasing trend during 2011-2021, the prevalence of SHT exposure was higher in schoolchildren from families with tobacco product users than in schoolchildren from families without tobacco product users, regardless of the type of tobacco product.

## INTRODUCTION

There is accumulating scientific evidence that secondhand smoke exposure has detrimental effects on the health of nonsmokers, including premature death [1]. Secondhand smoke exposure is one of the most important adverse exposures in childhood, with children accounting for 28% of deaths from secondhand smoke worldwide [2]. Children are particularly susceptible to secondhand smoke exposure because of their high breathing rate and large lung surface area [3]. Middle-ear disease, asthma, wheeze, cough, bronchitis, pneumonia, and pulmonary dysfunction are common in children with secondhand smoke exposure [4,5]. Recently, it has been reported that secondhand smoke exposure in childhood is also associated with an increased risk of respiratory disease-related mortality, coronary heart disease mortality, and pancreatic cancer mortality in adulthood [6–8].

Japan has been a major market for heated tobacco products (HTPs) since November 2014, when IQOS, one of the most popular brands of HTPs, was launched in Japan ahead of the world. The number of HTP users has increased rapidly in recent years, with younger people in the child-rearing generation more likely to use HTPs [9]. Now, HTPs are the second most used tobacco product after traditional combustible cigarettes (CCs), with an estimated users of 19.4% for CCs and 11.8% for HTPs in 2022 [10]. Against the background of the rapid spread of HTPs in Japan, our recent study estimated that about one in ten people were already exposed to second-hand aerosols from HTPs as of 2020 [11].

The emergence of HTPs has complicated the issue of secondhand tobacco (SHT) exposure, such as secondhand exposure to CC smoke and HTP-generated aerosols. Although the aerosol generated from HTPs contains significantly lower amounts of harmful and potentially harmful constituents than those in CC smoke [12–15], there is currently no evidence to indicate that HTPs are less harmful than conventional tobacco products [16,17]. In fact, a recent cross-sectional study in Japan reported the association between secondhand-aerosol exposure from HTPs and asthma attacks/asthma-like symptoms and persistent cough among current non-users [18]. In other words, it is important to monitor aerosol exposure from HTPs for not only HTP users but also bystanders including children.

Although assessing the extent of tobacco smoke exposure based on self-report may be subject to response bias [19], biomonitoring of urinary levels of nicotine metabolites, including cotinine, is an accurate and widely used means of assessing SHT exposure [20–22]. One of them, urinary levels of cotinine, are reported to be significantly higher in nonsmokers exposed to SHT products, regardless of tobacco type (CCs or HTPs) [22]. There are previous studies focusing on trends in SHT smoke exposure as verified by urinary cotinine levels in non-smoking adults [23,24], but, to our knowledge, there are no studies in schoolchildren. In addition, when self-reported SHT exposure is included, there have been reports for junior and high school students [25–27], but none for elementary school students. Therefore, the main goals of this study were to evaluate the trends in the prevalence of SHT exposure as measured by urinary cotinine levels among Japanese elementary school children from 2011 to 2021 and to examine the association between SHT exposure and household tobacco product use.

## METHODS

### Study population and setting

This repeated cross-sectional study was based on data of the survey that was annually conducted in Kumagaya City from 2011 to 2021 by the Education Board to investigate the prevalence of SHT exposure, such as secondhand exposure to CC smoke and HTP-generated aerosols, among school-aged children. Kumagaya City is located in Saitama Prefecture, Kanto Region, Japan, and had a residential population of approximately 200,000. All the children in fourth grade (aged 10 years) in Kumagaya City were recruited to participate in the survey through the elementary schools. In the survey, children were measured urinary cotinine levels to objectively assess whether they were exposed to SHT. In addition, the parents or guardians were offered to answer a questionnaire that included questions on the tobacco product use status of the household members who lived together. From 17,596 children with the information about questionnaire results, we extracted 15,927 children with the information about urinary cotinine levels as the analytical sample. The number of participants who did not have the information about urinary cotinine levels by survey year from 2011 to 2021 are shown in Supplemental Table 1.

This study included the two analyses (analyses 1 and 2) with different purposes. Analysis 1 investigated the annual trend of the prevalence of SHT exposure evaluated by the urinary cotinine levels. Analysis 2 assessed the prevalence of SHT exposure evaluated by the urinary cotinine levels according to the household tobacco use status. While analysis 1 was based on the data from 2011 to 2021, analysis 2 used the data from 2018 to 2021 since the question about HTP use status was included in the questionnaire from 2018.

### Urinary cotinine measurement and definition of SHT exposure

The children were instructed to collect the first-morning urine samples in a sterile container provided in advance. The samples were sent from their schools to the designed laboratory (Cosmic Corporation Co., Ltd., Tokyo, Japan; BML Inc., Tokyo, Japan) within the day. Then, the urinary cotinine levels were measured using a monoclonal antibody-based competitive enzyme-linked immunosorbent assay (ELISA). In the method, the limit of quantitation for urinary cotinine levels was 1.3 ng/ml. According to the definition by the Japan Society for Tobacco Control [28] and the definitions used in previous study [29], we classified the children with urinary cotinine levels ≥5.0 ng/ml into the cotinine verified SHT exposed group. In the sensitivity analysis, we used a different cut-off (≥3.0 ng/ml) in the definition of SHT exposure to assess the potential usefulness of the cut-off in future studies.

### Household tobacco use

The household tobacco use status was defined by whether there was at least one household member who used tobacco products at their home. In the questionnaire, household members were asked about the number of CCs and HTPs used on the day before the children collected the urine sample. We regarded the member who used at least one as a current CCs or HTPs user in this study according to the definitions in previous studies [10,30,31]. Subsequently, we defined the household tobacco use status by aggregating the household members’ tobacco product use status at the household level into the following four categories: none used, only CCs, only HTPs, and CCs and HTPs. Here, “only CCs” denotes that the children had at least one household member who was a CC user and did not have members who were HTP users.

### Statistical analysis

We presented the descriptive characteristics of the analytical sample by the SHT exposure status. As analysis 1, we constructed a scatter plot of the urinary cotinine levels and a forest plot of the prevalence of SHT exposure by survey year from 2011 to 2021. In the forest plot, we fitted an approximated curve with 95% confidence intervals for the mean prevalence of SHT exposure over the period. As analysis 2, we constructed a scatter plot of the urinary cotinine levels and a forest plot of the prevalence of SHT exposure by the household tobacco use status. In addition, we assessed the differences in the mean prevalence of SHT exposure by the household tobacco use status using t-tests.

As additional analyses, we assessed the annual trends from 2018 to 2021 on the prevalence of CC or HTP users among the children’s household members who were tobacco product users. In addition, we assessed the number of tobacco product users among the household members by survey year from 2011 to 2021. All analyses were conducted using Stata version 17.0 (Stata Corp, College Station, TX). Two-sided *P*-values <0.05 were considered statistically significant in all cases. The reporting of this study followed the Strengthening the Reporting of Observational Studies in Epidemiology (STROBE) guidelines.

### Ethical considerations

This study was approved by the Ethics Committee of Tohoku University Graduate School of Dentistry (approval number: 34020). This study was based on the de-identifiable data approved to be used and provided by the Education Board of Kumagaya City. Informed consent was obtained from the children’s parents or guardians by the Education Board of Kumagaya City.

## RESULTS

Table 1 presents the characteristics of the analytical sample. Among 15,927 participants (male, 8,250; female, 7,677), 1,887 (11.8%) were exposed to SHT assessed by the urinary cotinine levels. There were 7,999 (50.2%) participants who had at least one tobacco product user in their household. In addition, among the participants who were exposed to SHT, 1,727 (91.5%) had at least one tobacco product user in their household. Among the participants’ household members, the prevalence of tobacco product users was 41.3% in father, 17.9% in mother, 2.3% in grandparents, 0.4% in siblings, and 0.8% in others.

**Table 1.**
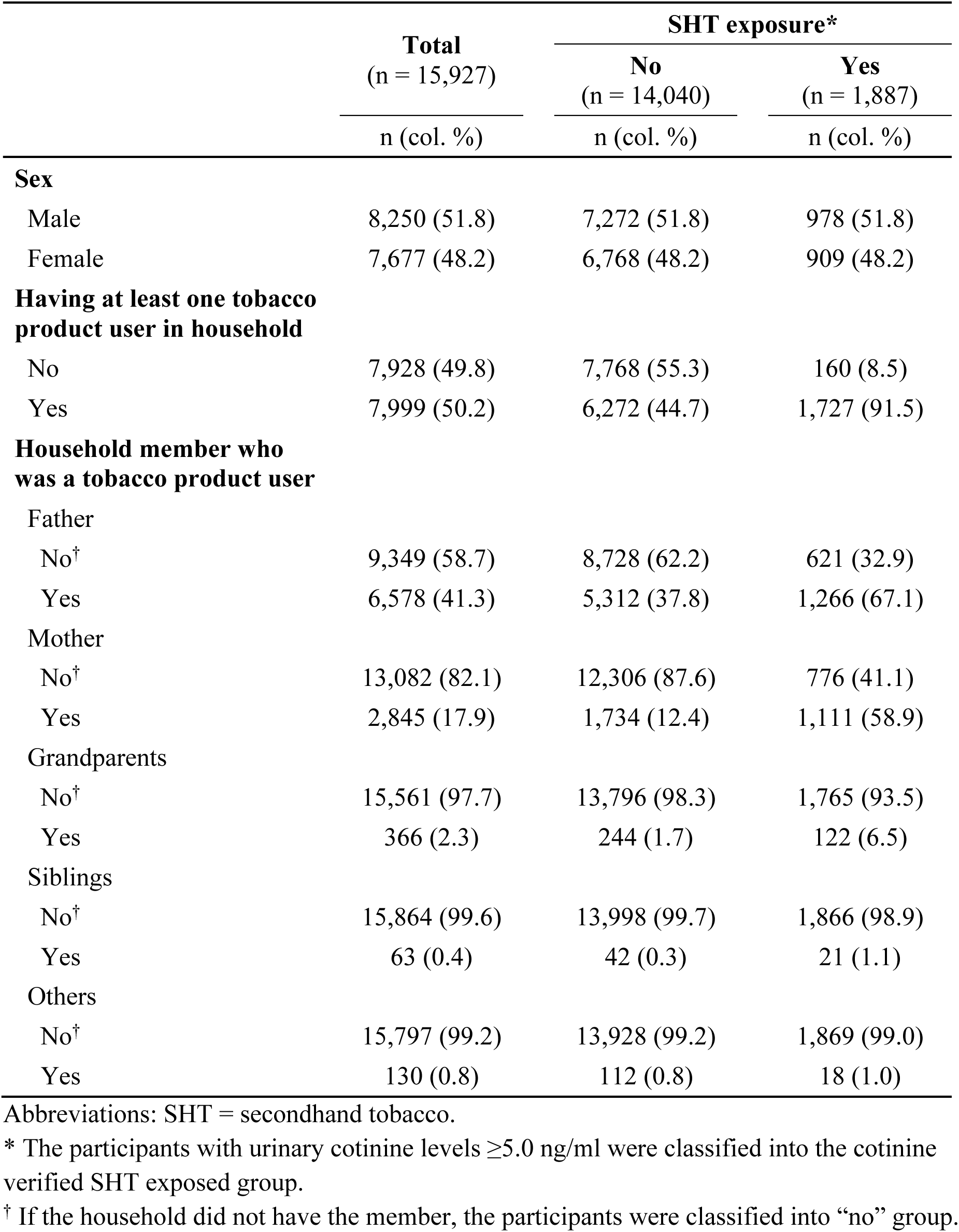
Characteristics of analytical sample according to SHT exposure status.

|  | **Total**<br>(n = 15,927) | **SHT exposure\*** |  |
| --- | --- | --- | --- |
|  |  | **No**<br>(n = 14,040) | **Yes**<br>(n = 1,887) |
|  | n (col. %) | n (col. %) | n (col. %) |
| **Sex** |  |  |  |
| Male | 8,250 (51.8) | 7,272 (51.8) | 978 (51.8) |
| Female | 7,677 (48.2) | 6,768 (48.2) | 909 (48.2) |
| **Having at least one tobacco product user in household** |  |  |  |
| No | 7,928 (49.8) | 7,768 (55.3) | 160 (8.5) |
| Yes | 7,999 (50.2) | 6,272 (44.7) | 1,727 (91.5) |
| **Household member who was a tobacco product user** |  |  |  |
| Father |  |  |  |
| No<sup>†</sup> | 9,349 (58.7) | 8,728 (62.2) | 621 (32.9) |
| Yes | 6,578 (41.3) | 5,312 (37.8) | 1,266 (67.1) |
| Mother |  |  |  |
| No<sup>†</sup> | 13,082 (82.1) | 12,306 (87.6) | 776 (41.1) |
| Yes | 2,845 (17.9) | 1,734 (12.4) | 1,111 (58.9) |
| Grandparents |  |  |  |
| No<sup>†</sup> | 15,561 (97.7) | 13,796 (98.3) | 1,765 (93.5) |
| Yes | 366 (2.3) | 244 (1.7) | 122 (6.5) |
| Siblings |  |  |  |
| No<sup>†</sup> | 15,864 (99.6) | 13,998 (99.7) | 1,866 (98.9) |
| Yes | 63 (0.4) | 42 (0.3) | 21 (1.1) |
| Others |  |  |  |
| No<sup>†</sup> | 15,797 (99.2) | 13,928 (99.2) | 1,869 (99.0) |
| Yes | 130 (0.8) | 112 (0.8) | 18 (1.0) |

Abbreviations: SHT = secondhand tobacco.
\* The participants with urinary cotinine levels $\geq 5.0$ ng/ml were classified into the cotinine verified SHT exposed group.
<sup>†</sup> If the household did not have the member, the participants were classified into “no” group.

Figure 1 A is the scatter plot of urinary cotinine levels by survey year. The figure indicated a declining trend in the number of participants with urinary cotinine levels ≥5.0 ng/ml from 2011 to 2021. In addition, there was a smaller number of participants with a high level of urinary cotinine (≥10.0 ng/ml) in later years. Figure 1 B is the forest plot of the prevalence of SHT exposure by survey year. The figure showed a declining trend in the prevalence of SHT exposure from 2011 to 2021. The prevalence declined from around 20% in 2011–2013 to 5% in 2018–2021.

**Figure 1.**
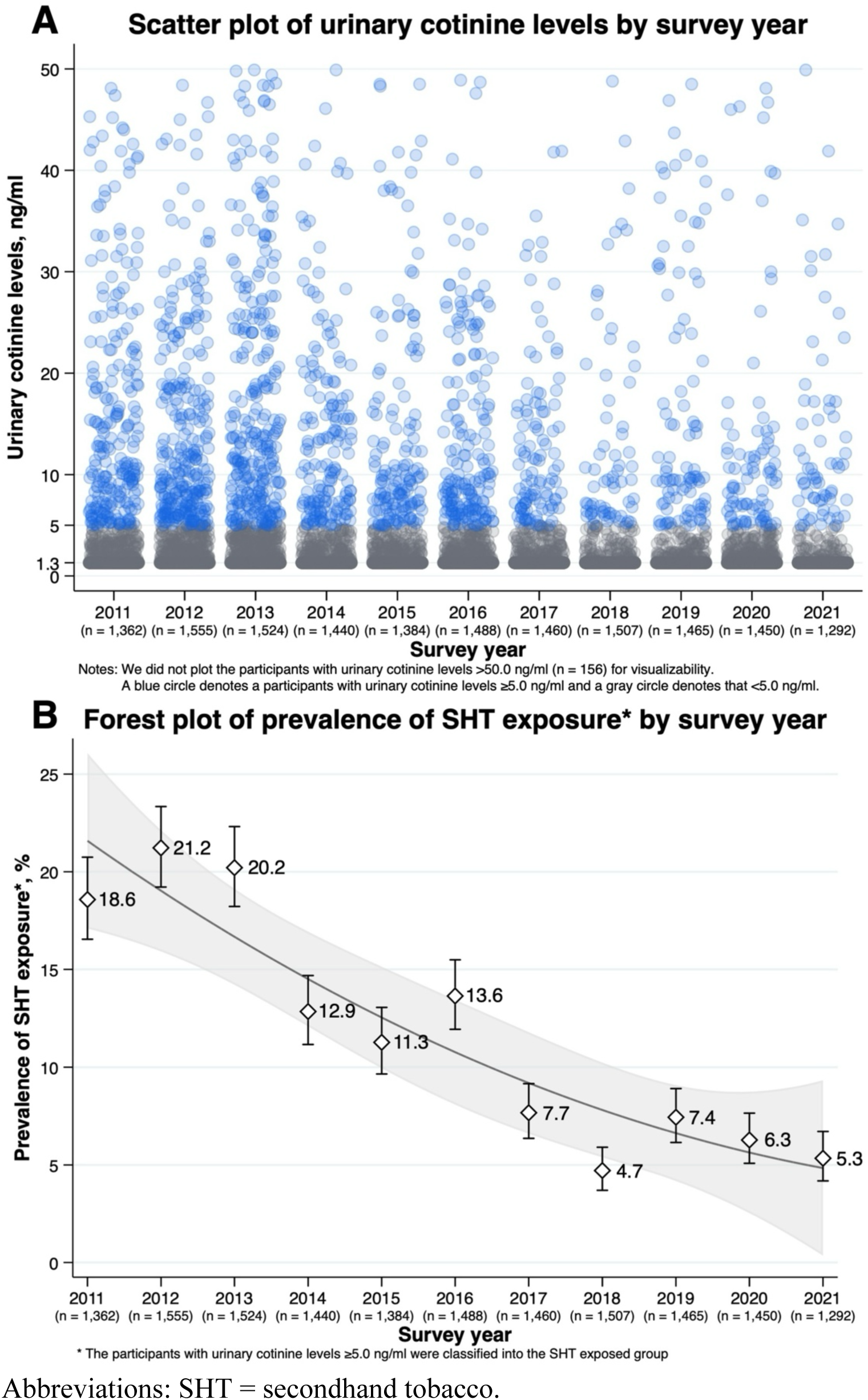
(A) Scatter plot of urinary cotinine levels by survey year (B) forest plot of prevalence of SHT exposure by survey year.

Figure 2 A is the scatter plot of urinary cotinine levels by household tobacco use status. Among the participants, 3,894 (68.1%) did not have tobacco product users, 941 (16.5%) had only CC users, 705 (12.3%) had only HTP users, and 174 (3.0%) had both CC users and HTP users in their household. In the figure, there were several fractions of participants with urinary cotinine levels ≥5.0 ng/ml especially in the household that had CC users. Figure 2 B is the forest plot of the prevalence of SHT exposure by household tobacco use status. Compared with the participants who did not have tobacco product users in their household, the prevalence of SHT exposure was higher in those who had only CC users (*P*<0.001), only HTP users (*P*=0.001), and both CC users and HTP users (*P*<0.001). In addition, compared with the participants who had only HTP users in their household, the prevalence of SHT exposure was higher in those who had only CC users (*P*<0.001) and both CC users and HTP users (*P*<0.001). On the other hand, there was no evidence on the difference in the prevalence of SHT exposure between the participants who had only CC users and both CC users and HTP users (*P*=0.177).

**Figure 2.**
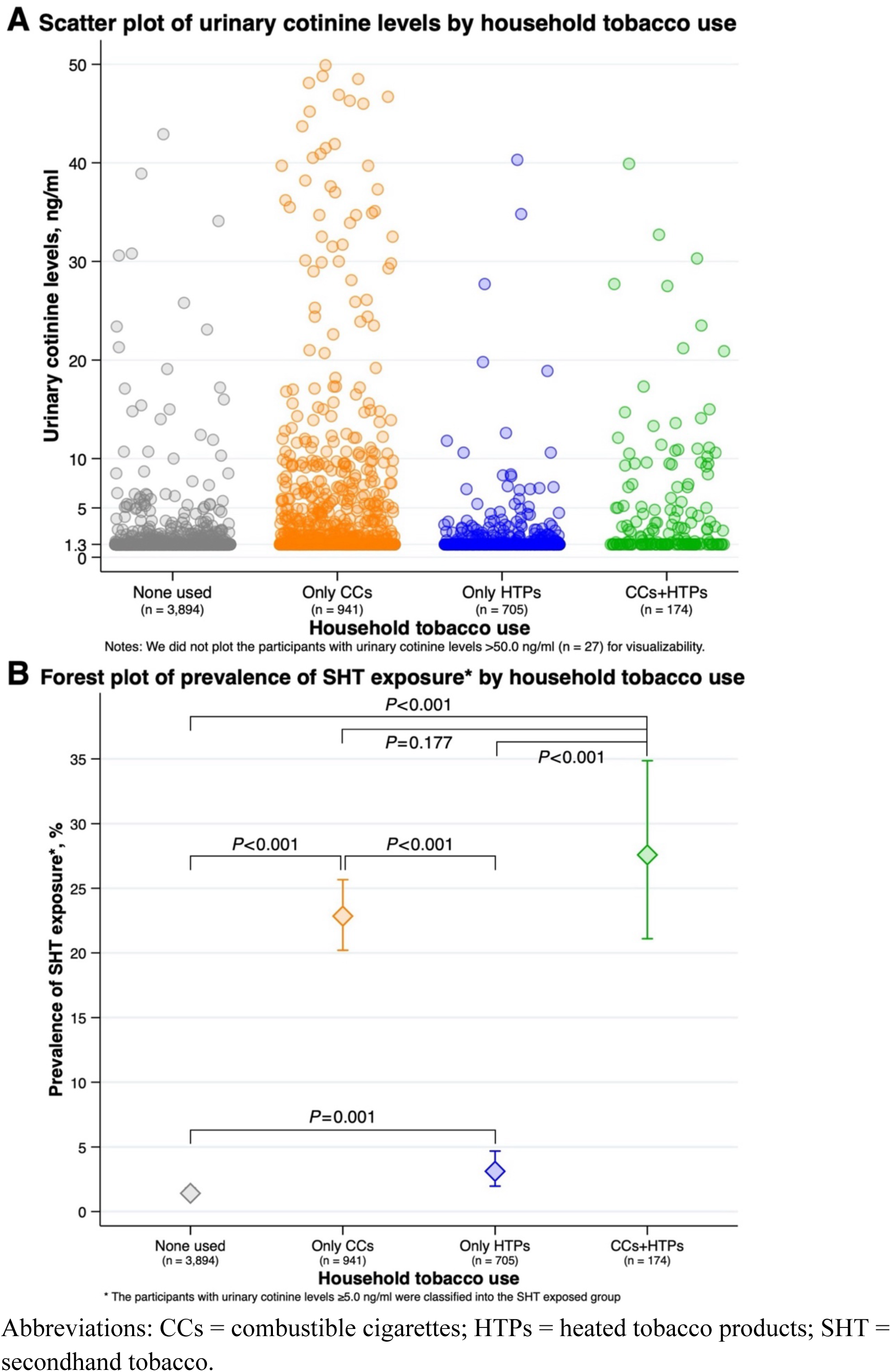
(A) Scatter plot of urinary cotinine levels by household tobacco use status (B) forest plot of prevalence of SHT exposure by household tobacco use status.

Largely similar results were observed in the sensitivity analysis that used a different cut-off (urinary cotinine levels ≥3.0 ng/ml) in the definition of SHT exposure. The prevalence of SHT exposure was on a declining trend from 2011 to 2021, and the prevalence declined from around 25% in 2011–2013 to 10% in 2018–2021 (Supplemental Figure 1). In addition, the prevalence of SHT exposure was higher in the participants with CC or HTP users in their households than those who did not have tobacco product users. Furthermore, the prevalence of SHT exposure was higher in the participants with CC users in their households than those with HTP users (Supplemental Figure 2). In the additional analysis, there was an increasing trend in the prevalence of HTP users among the tobacco product users in the household, while the prevalence of CC users was on a declining trend (Supplemental Figure 3). In addition, there was a declining trend in the prevalence of tobacco product users. The prevalence of tobacco product users declined from 46.5% in 2011 to 35.8% in 2021 in fathers and from 19.2% in 2011 to 12.6% in 2021 in mothers (Supplemental Table 2).

## DISCUSSION

SHT exposure, as confirmed by urinary cotinine levels, among Japanese elementary school students has decreased by a quarter over the past 11 years, from about 20% in the early 2010s to about 5% in the early 2020s. On the other hand, significant differences existed in urinary cotinine levels among elementary school students by household tobacco use status. Compared to elementary school children without tobacco product users in the household, the prevalence of SHT exposure was significantly higher in households with tobacco product users, regardless of the type of tobacco product. This suggests that even in households where only HTP was used, children in the household were exposed to SHT exposure similar to conventional secondhand CC smoke.

A previous study [27] observed a decrease of SHT exposure among young people in middle adolescence in Japan in the similar period (2008–2017) to our study, but the current situation of SHT exposure among the younger population than the study remained unknow. Here, our study found a continuous decrease of SHT exposure among young people in early adolescence from 2011 to 2021, which extends the understanding of current situation of SHT exposure among children in Japan. Considering that smokers have been decreasing in Japan (as supported by the results in Supplemental Table 2), the decrease of SHT exposure among children may be explained by the decrease of chances that smokers would be around children. In addition, the Revised Health Promotion Act prohibits tobacco use in public places from April 2020 in Japan [32]; hence, SHT exposure is expected to further decrease in the coming years. However, when a similar act, which prohibit tobacco use in indoor public places, was implemented in Spain, it did not work to reduce SHT exposure to children [33]. Therefore, we consider that the prevalence of SHT exposure among children should be continuously monitored to evaluate the effectiveness of Revised Health Promotion Act and provide feedback to refine the act.

This study observed that the prevalence of SHT exposure was lower among children with only HTP users in their household than those with CC users. This may reflect that HTPs contain the same or less amount of nicotine with CCs per stick [34]. However, compared with CCs, HTPs are reported to contain more amount of some harmful and potentially harmful constituents [35], including those whose long-term effects on health are unknown. In these contexts, it would be important to call more attention to evaluate SHT exposure using a biomarker whether it adequately capture the exposure. Our results in Supplemental Figure 1 and 2 may encourage the use of more strict cut-off of urinary cotinine levels in the definition of SHT exposure to account for the use of a variety of tobacco products nowadays.

The strength of this study is that it is the first large-scale examination of trends in SHT exposure among Japanese elementary school students over a 10-year period using urinary cotinine testing and its association with the household use of tobacco products including HTS. On the other hand, this study has several limitations. First, this study was based on data from students attending elementary school in a single municipality. Therefore, our findings should be interpreted with caution, especially when generalizing to the same age group not attending school or to elementary school students in other municipalities. However, the most recent Ministry of Education, Culture, Sports, Science and Technology (MEXT) survey reported that the percentage of students not attending school in Japanese elementary schools is 1.7% [36], and almost all children of this age group are attending school. Further research using data from multiple municipalities is needed to assess the generalizability of the findings. Second, although this study focused on the presence of tobacco product users in households as one of the causes of SHT exposure, the possibility that children are exposed to SHT outside the home (e.g., on the way to school) cannot be denied. However, our study subjects were 10-year-old elementary school children, who are expected to spend most of their time at home [37]. In addition, with the full enforcement of the revised Health Promotion Law in Japan in April 2020, smoking in public places, including around schools, is basically banned, and we believe that SHT exposure outside the home is quite unlikely to affect the results of this study.

In conclusion, this study revealed a consistent downward trend in the prevalence of SHT exposure among elementary school students in one Japanese municipality between 2011 and 2021. However, the prevalence of SHT exposure was found to be higher among elementary school students in households with tobacco product users than among those without tobacco product users in households, regardless of the type of tobacco product. These findings suggest that using of tobacco products, including HTPs, in households with children should be considered for legal regulation in order to avoid adverse health effects in populations that cannot choose whether or not to accept the risks derived from SHT exposure, such as children in the home.

## Appendix

Supplementary material can be found in the online version of the journal.

## Acknowledgments

We are grateful to the participants of this study for their valuable contributions.

## Author contribution

YT, KT, and TT were involved in the study design, data analysis, and data interpretation. TT collected and checked data. YT and KT wrote the first draft of the manuscript. KT and TT supervised and administrated the study. TT acquired funding. All authors gave their final approval and agreed to be accountable for all aspects of the work.

## Data availability statement

All data used in this study are not publicly available due to ethical or legal restrictions.

