## Supplemental file for "Secondhand tobacco exposure assessed using urinary cotinine among 10 years children in Japan: A 11-years repeated cross-sectional study"

**Title:**

**Supplemental Table 1.** Number of participants who did not have urinary cotinine information by survey year (2011–2021).

| **Survey year** | **Having urinary cotinine information**, n (row %) | **Not having urinary cotinine information**, n (row %) | **Total** |
|---|---|---|---|
| 2011 | 1,362 (99.3) | 9 (0.7) | 1,371 (100.0) |
| 2012 | 1,555 (87.3) | 227 (12.7) | 1,782 (100.0) |
| 2013 | 1,524 (89.3) | 183 (10.7) | 1,707 (100.0) |
| 2014 | 1,440 (89.1) | 176 (10.9) | 1,616 (100.0) |
| 2015 | 1,384 (87.9) | 190 (12.1) | 1,574 (100.0) |
| 2016 | 1,488 (90.2) | 161 (9.8) | 1,649 (100.0) |
| 2017 | 1,460 (87.2) | 215 (12.8) | 1,675 (100.0) |
| 2018 | 1,507 (93.8) | 100 (6.2) | 1,607 (100.0) |
| 2019 | 1,465 (93.0) | 111 (7.0) | 1,576 (100.0) |
| 2020 | 1,450 (91.2) | 140 (8.8) | 1,590 (100.0) |
| 2021 | 1,292 (89.2) | 157 (10.8) | 1,449 (100.0) |
| Total | 15,927 (90.5) | 1,669 (9.5) | 17,596 (100.0) |

**Supplemental Table 2.** Number of tobacco product users among household members by survey year (2011–2021).

| **Survey year** | **Father**, n (row %) | **Mother**, n (row %) | **Grandparents**, n (row %) | **Siblings**, n (row %) | **Others**, n (row %) | **Total** |
|---|---|---|---|---|---|---|
| 2011 | 633 (46.5) | 262 (19.2) | 39 (2.9) | 0 (0.0) | 3 (0.2) | 1,362 |
| 2012 | 679 (43.7) | 300 (19.3) | 18 (1.2) | 5 (0.3) | 10 (0.6) | 1,555 |
| 2013 | 690 (45.3) | 276 (18.1) | 35 (2.3) | 1 (0.1) | 5 (0.3) | 1,524 |
| 2014 | 655 (45.5) | 254 (17.6) | 92 (6.4) | 4 (0.3) | 25 (1.7) | 1,440 |
| 2015 | 590 (42.6) | 364 (26.3) | 0 (0.0) | 6 (0.4) | 14 (1.0) | 1,384 |
| 2016 | 625 (42.0) | 356 (23.9) | 0 (0.0) | 10 (0.7) | 15 (1.0) | 1,488 |
| 2017 | 576 (39.5) | 301 (20.6) | 0 (0.0) | 12 (0.8) | 18 (1.2) | 1,460 |
| 2018 | 570 (37.8) | 176 (11.7) | 55 (3.7) | 6 (0.4) | 10 (0.7) | 1,507 |
| 2019 | 570 (38.9) | 213 (14.5) | 44 (3.0) | 4 (0.3) | 13 (0.9) | 1,465 |
| 2020 | 528 (36.4) | 180 (12.4) | 41 (2.8) | 8 (0.6) | 8 (0.6) | 1,450 |
| 2021 | 462 (35.8) | 163 (12.6) | 42 (3.3) | 7 (0.5) | 9 (0.7) | 1,292 |
| Total | 6,578 (41.3) | 2,845 (17.9) | 366 (2.3) | 63 (0.4) | 130 (0.8) | 15,927 |

**Supplemental Figure 1.** Forest plot of prevalence of SHT exposure by survey year using a different cut-off (≥3.0 ng/ml) in definition of SHT exposure.

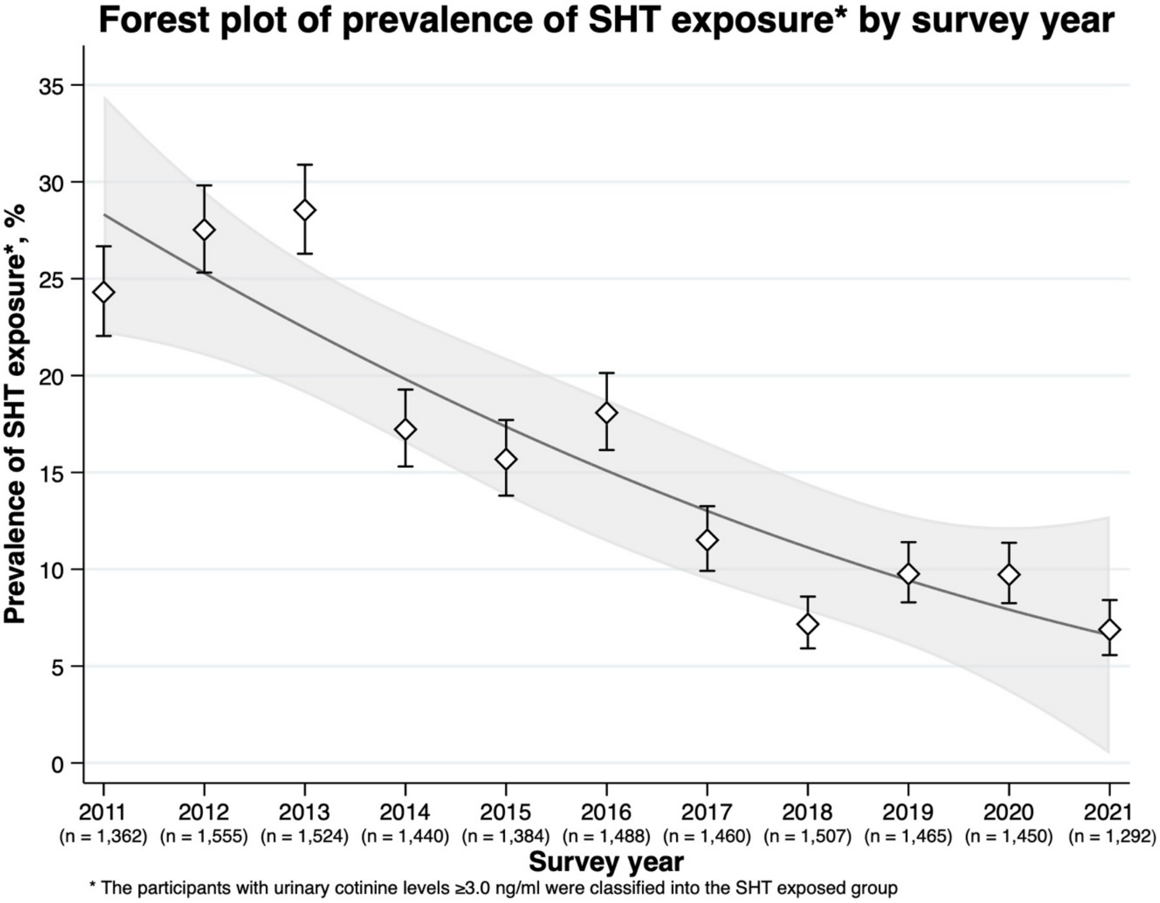

Abbreviations: SHT = secondhand tobacco.

**Supplemental Figure 2.** Forest plot of prevalence of SHT exposure by household tobacco use using a different cut-off (≥3.0 ng/ml) in definition of SHT exposure.

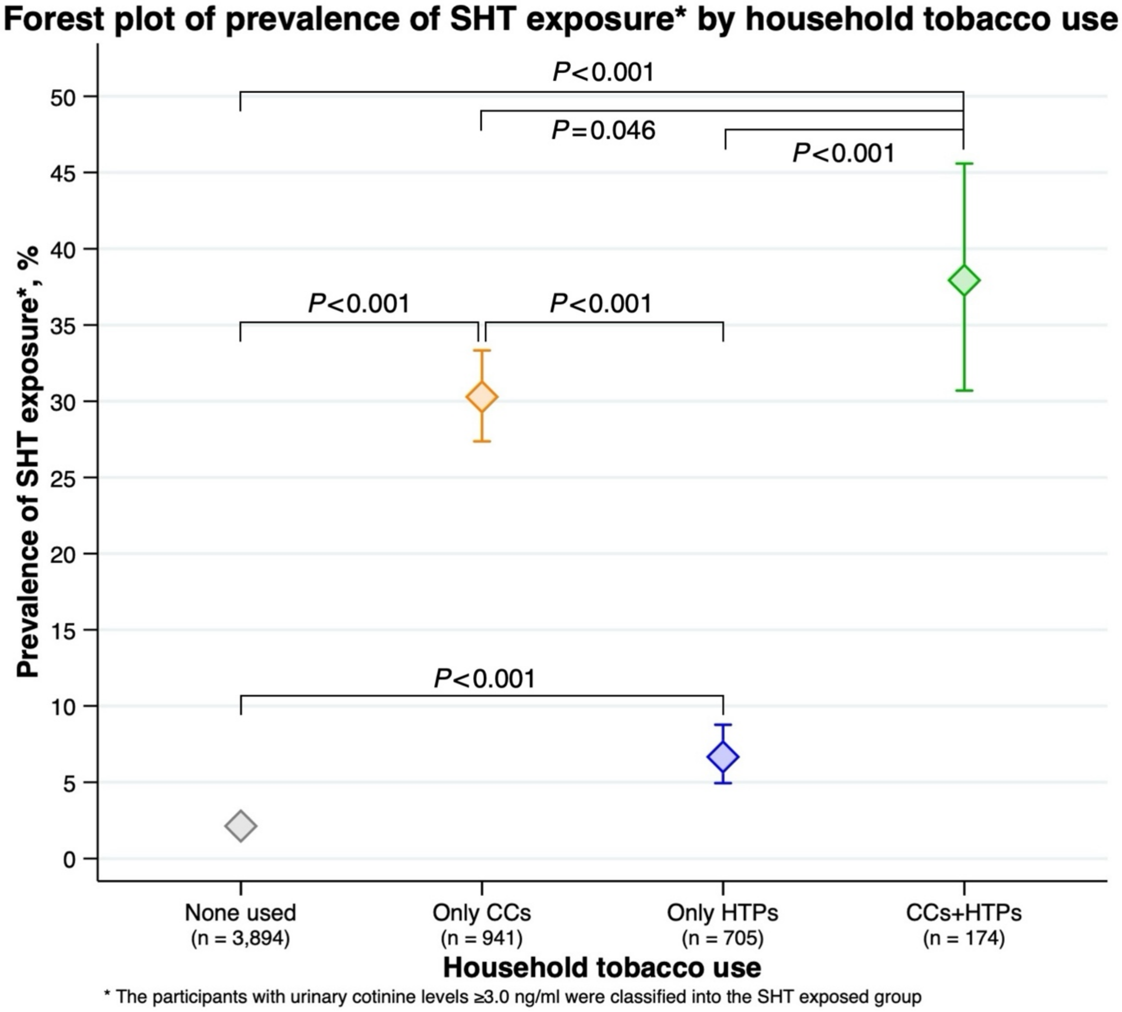

Abbreviations: CCs = combustible cigarettes; HTPs = heated tobacco products; SHT = secondhand tobacco.

**Supplemental Figure 3.** Annual trends from 2018 to 2021 on prevalence of CCs or HTPs users among tobacco product users in household.

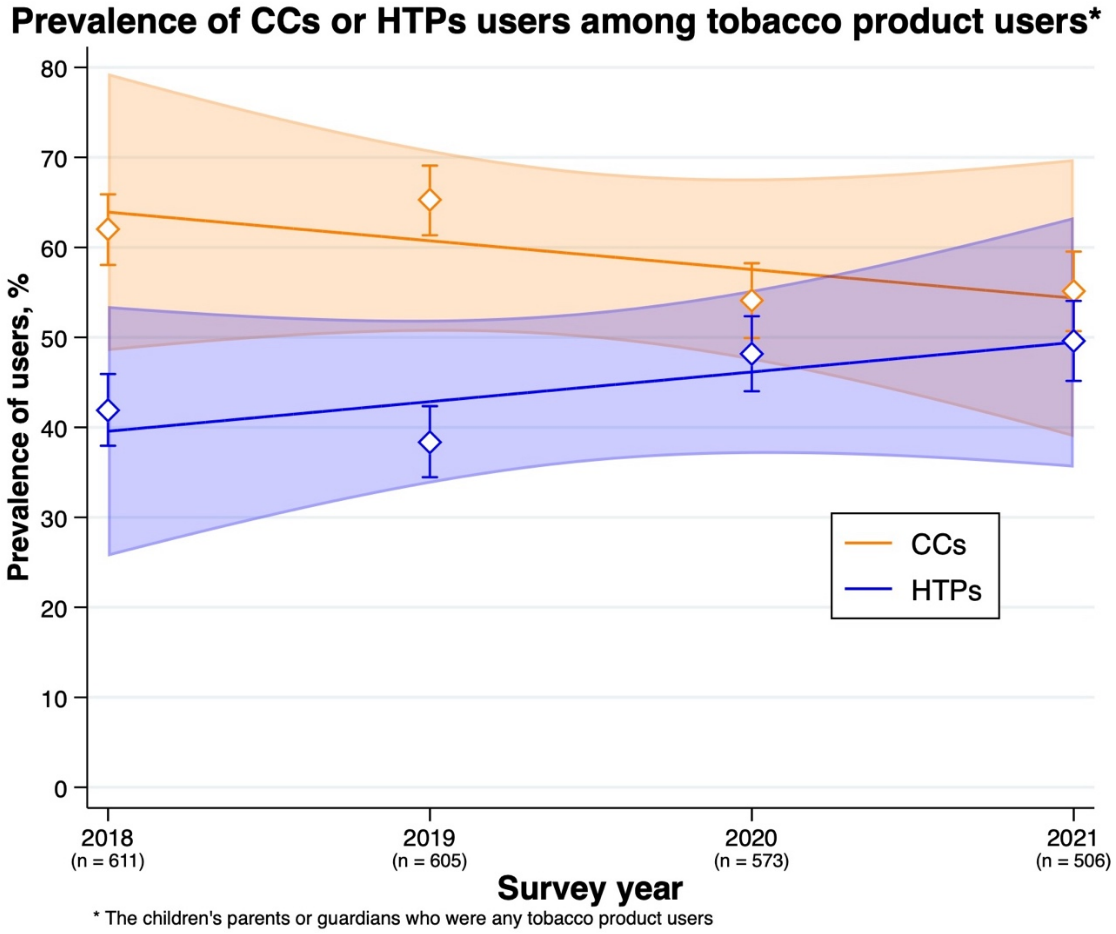

Abbreviations: CCs = combustible cigarettes; HTPs = heated tobacco products.
